# Plasma Short-Chain Fatty Acids and Hypertensive Disorders of Pregnancy: Results from a Nested Case-Control Study and Two-sample Mendelian Randomization Analysis

**DOI:** 10.1101/2024.08.14.24312020

**Authors:** Dandan Wang, Lei Wu, Pei Feng, Yexiu Sun, Di Wu, He Xu, Hao Peng, Hongmei Li

**Author notes:** These authors contributed equally to this work and should be considered co-first authors (Dandan Wang, Lei Wu, Pei Feng). Corresponding author: Hao Peng, MD, PhD; Hongmei Li, MD, PhD, **Contact Info**: Hao Peng, MD, PhD, Department of Epidemiology and Biostatistics, School of Public Health, Suzhou Medical College of Soochow University. 199 Renai Road, Industrial Park District, Suzhou 215123, PR China., Hongmei Li, MD, PhD, Department of Epidemiology and Biostatistics, School of Public Health, Suzhou Medical College of Soochow University. 199 Renai Road, Industrial Park District, Suzhou 215123, PR China.

## Abstract

**Background:** The temporal relationship between Short-chain fatty acids (SCFAs) and hypertensive disorders of pregnancy (HDP) is unclear. This study aimed to examine the temporal and probable causal relationship between them.

**Methods:** A nested case-control study including 387 pairs of cases with HDP and healthy controls was conducted. Seven SCFAs levels in plasma samples drawn at 16-20 gestational weeks before HDP were assayed by GC/MS. The individual and joint associations of SCFAs with HDP were examined by logistic regression and weighted quantile sum (WQS) regression, respectively, followed by two-sample bidirectional Mendelian randomization (MR) analysis to test the underlying causality.

**Results:** The univariate model found each interquartile increase in plasma valerate was associated with a 32.1% (OR=0.679, 95%CI=0.546-0.844) reduction in the risk of HDP, a 29.4%(OR=0.706, 95%CI=0.548-0.910) reduction in the risk of gestational hypertension (GH) and a 39.1% (OR=0.609, 95%CI=0.397-0.935) reduction in the risk of preeclampsia/chronic hypertension with superimposed preeclampsia (PE/CH-PE). However, after adjustment for covariates, valerate was only associated with the risk of HDP (OR=0.699, 95% CI=0.516-0.946). In addition, plasma isobutyrate and hexanoate were associated with lower risks of HDP and PE/CH-PE. Furthermore, SCFAs co-exposure could reduce the risks of HDP and GH. MR showed that plasma acetate (OR=0.784, 95%CI=0.64-0.962), valerate (OR=0.575, 95%CI=0.363-0.909) and isovalerate (OR=0.642, 95%CI=0.428-0.963) had protective causal effects on GH. Meanwhile, plasma acetate had protective causal effects on PE (OR=0.746, 95%CI=0.6-0.927).

**Conclusions:** The study suggested it is necessary to appropriately increase SCFAs levels during pregnancy to reduce the risk of HDP.

## Introduction

Hypertensive disorders of pregnancy (HDP), mainly comprised of gestational hypertension (GH) and preeclampsia (PE), are not only the leading cause of maternal and fetal mortality but also associated with a wide range of disorders for both mothers and infants in later life ^1,2^. It is the most common medical disorder occurring during pregnancy, complicating about 3%-10% of pregnancies and being increasingly prevalent globally, China in particular ^3,4^. Identification of more risk factors and biomarkers would undoubtedly improve the prevention and management of this debilitating disorder. The fundamental pathogenesis of HDP is inadequate adaptive morphological processes at the maternal and fetal interface ^4–6^. Recent evidence suggests that gut microbiota, a vast and complex collection of microorganisms, is engaged in the adaption of hosting women for their dynamic pregnancy ^7^. As the main metabolic end products of gut microbes, short-chain fatty acids (SCFAs) may participate in and could be candidate biomarkers for the development of HDP. Indeed, this hypothesis has been suggested by several studies. For example, SCFAs have been associated with blood pressure and original hypertension in adults without pregnancy ^8,9^. Animal experiments found that supplementation of butyrate could significantly reduce blood pressure in rats with PE ^10^. Deficiency of butyrate in the circulation was also observed in pregnant women with PE, compared with healthy controls ^11^. Circulating levels of other SCFAs, e.g., hexanoate, acetate, propionate, isobutyrate, and valerate were also differed between pregnant women with and without PE in previous studies, but with mixed results ^11,12^. These results were mainly from case-control studies and it is still unclear about the temporal relationship between SCFAs and HDP which is critical for causal inference. Therefore, this study aimed to investigate the causal relationship between SCFAs and the incidence of HDP.

## Methods

### Study design

We conducted a two-step design, specifically, a nested case-control study design was used to examine the temporal association between 7 SCFAs in plasma at 16-20 gestational weeks (gw) and HDP, followed by a two-sample bidirectional Mendelian Randomization (MR) analysis with web-based summarized data to explore the causal relationship between 3 certain SCFAs and HDP.

### Participants

Suzhou Maternal and Infant Cohort (SMIC) is a hospital-based prospective cohort study aiming to examine the effect of environmental factors during pregnancy on the health of pregnant women and their offspring. This study was initiated in 2019 by including singleton pregnant women aged over 18 years in Kunshan and Industrial Park, respectively. After providing written informed consent, all participants received questionnaires and were offered free physical examinations, blood cell analysis, and clinical biochemical tests using blood and urine specimens during the whole pregnancy, under the principle of voluntary acceptance. Antenatal visits were performed every four weeks before 28 gw, every two weeks from 28 to 36 gw, and every one week after 36 gw until delivery. The protocols of this study were approved by the Ethics Committee of Soochow University (NO. ECSU-2019000118).

Of the sub-cohort including 9447 participants who completed all antenatal examinations from Kunshan, 415 individuals developed HDP after 20 gw and 9032 participants remained free of HDP during the whole pregnancy. After further excluding participants with a history of heart disease, stroke, chronic kidney disease, or tumors, 403 cases of HDP and 403 age- and parity-matched healthy controls were randomly selected and their plasma samples obtained at 16-20 gw were used to measure SCFAs concentrations. After laboratory testing, 16 pairs who failed to detect SCFAs were excluded, and 387 pairs in which both cases and controls with available data on SCFAs were included in the current analysis. The flow chart for screening subjects is shown in **Supplementary Figure S1.**

### Measurement of blood pressure and the definition of HDP

Three consecutive sitting blood pressure (BP) measurements (with 30s in-between) were taken by trained staff using a digital BP measuring device (HBP-1320, Omron, Japan) at each prenatal visit, after resting for at least 30 min. The mean of the three measurements was used as the visit BP level. Cases of HDP in our study included GH, PE, and chronic hypertension with superimposed PE (CH-PE) ^5^. Participants who were free of hypertension before pregnancy but developed hypertension (BP≥140/90 mmHg) for the first time at 20 gw or later without proteinuria were diagnosed with GH. Those with GH complicated with proteinuria, maternal end-organ complication, or evidence of uteroplacental dysfunction were diagnosed with PE. Participants with hypertension before pregnancy but superimposed on PE were diagnosed with CH-PE. No participants suffered from eclampsia in our study.

### Measurement of plasma SCFAs

Blood samples were collected at 16-20 gw before the development of HDP and frozen at - 80 ℃ until laboratory assay. Plasma levels of seven SCFAs including acetate, propionate, butyrate, isobutyrate, valerate, isovalerate, and hexanoate were determined using GC/MS method detailed elsewhere ^13^. In brief, after SCFAs extraction with butanol, the preprocessed samples were injected into and transported through the chromatographic column (Agilent HP-INNOWAX) by the helium carrier using gas chromatography (Thermo Trace 1310). Meanwhile, mass spectrometry (Thermo ISQ LT) was used to detect the seven compounds using electrospray ionization and selected ion monitoring (SIM).

### Collection of conventional risk factors

Data on age, education level, parity, and family history of hypertension and diabetes were collected through questionnaires by medical staff. Body weight and height were measured with the participants wearing light clothes and no shoes. Body mass index (BMI) was calculated using the formula of the weight in kilograms divided by the square of height in meters (kg/m^2^). Fasting plasma glucose and hemoglobin were measured by standard laboratory methods.

### Statistical analysis

To carefully delineate the role of SCFAs in the development of HDP, we first examined the associations between SCFAs and HDP using original data with a nested case-control study design, followed by a two-sample MR analysis using summarized data. All data analyses were performed using SAS 9.4 software (SAS Institute Inc. Cary, NC) and R software (version 4.1.2) with a significance level at a *P* value less than 0.05.

#### Case-control analysis

The baseline characteristics of study participants were presented in HDP cases and their matched healthy controls, respectively. To examine the associations between SCFAs and HDP, we constructed a conditional logistic regression model in which HDP (y/n) was the dependent variable and each SCFA (per quartile increment) was the independent variable, adjusting for maternal education level, BMI, systolic BP, hemoglobin, and fasting plasma glucose at the first antenatal visit. Due to the seven compounds assayed participating in HDP through similar pathways and interplaying with each other, we applied weighted quantile sum (WQS) regression to examine their joint association with HDP by constructing a weighted index to estimate the mixture exposure level of SCFAs. The WQS regression model was performed using the R package “gWQS”. The associations between SCFAs and specific subtypes of HDP were similarly examined.

#### Two-sample MR analysis

To delineate the probable causality underlying the associations between SCFAs and HDP, we performed a two-sample bidirectional MR analysis. The summarized genome-wide association study (GWAS) data for SCFAs, GH, and PE were obtained from publicly available databases. The SCFAs (acetate, valerate, and isovalerate) data were derived from the Estonian Genome Center of the University of Tartu Cohort, Finnish Twin Cohort, Helsinki Birth Cohort Study, Cooperative Health Research in the Region of Augsburg and so on ^14,15^. Data on GH and PE were selected from the Finngen consortium GWAS data including 209980 European-ancestry participants (https://www.finngen.fi/en). In the MR analysis, independent single nucleotide polymorphisms (SNPs) associated with SCFAs, GH, and PE (*P* < 5×10^-5^, linkage disequilibrium [LD] r^2^ < 0.001, F>10) were selected as instrumental variables (IVs). Given that obesity and diabetes are the high-impact confounders, SNPs associated with BMI and diabetes mellitus (*P*<10^-5^) were excluded (http://www.phenoscanner.medschl.cam.ac.uk/). The causal effect was estimated by inverse variance weighted (IVW), weighted median (WM), and MR-Egger regression ^16^. The heterogeneity of the IVs was tested using the Cochran Q test, and a significance threshold for evidence of heterogeneity was set at *P* <0.05. If heterogeneity existed, random-effect IVW models were used. Otherwise, fixed-effect IVW models were used. Potential pleiotropy in the IVs was examined by MR Egger. Leave-one-out approach was applied to test whether the causal estimate was biased by individual instrumental SNP.

## Results

### Characteristics of study participants

A total of 387 pairs of cases with HDP and age- and parity-matched healthy controls were included in the current study (median aged 28 years). Their clinical characteristics are shown in **Table 1**. Compared with healthy controls, cases with HDP were more likely to be less educated and have a family history of hypertension, gestational diabetes, overweight or obesity, and higher levels of FPG, hemoglobin, and BP at the first antenatal visit (all *P*<0.05).

**Table 1.** Clinical characteristics of study participants according to HDP.

| Variables | Case (*n*=387) | Control (*n*=387) | *P* |
| --- | --- | --- | --- |
| Maternal age, years | 28 (25 - 31) | 28 (25 - 31) | 0.930 |
| Parity, *times* | 2(1-3) | 2(1-3) | 0.350 |
| Education level, *n* (%) |  |  | <0.001 |
| ≤ Middle school | 111 (27.54) | 71 (17.62) |  |
| High school | 104 (25.81) | 82 (20.35) |  |
| ≥ College | 188 (46.65) | 250 (62.03) |  |
| Family history of hypertension (*n*, %) | 25 (6.46) | 0 (0.00) | <0.001 |
| Family history of diabetes (*n*, %) | 4 (1.03) | 3 (0.78) | 0.704 |
| Gestational diabetes (*n*, %) | 64(16.54) | 1(0.26) | <0.001 |
| Indicators at the first antenatal visit |  |  |  |
| BMI (*n*, %) |  |  | <0.001 |
| ≤23.9 kg/m<sup>2</sup> | 209(54.0) | 316(81.7) |  |
| 24.0 to 27.9 kg/m<sup>2</sup> | 106(27.4) | 57(14.7) |  |
| ≥28.0 kg/m<sup>2</sup> | 72(18.6) | 14(3.6) |  |
| Fasting plasma glucose (mmol/L), (M, Q<sub>L</sub>-Q<sub>U</sub>) | 4.64 (4.38-4.95) | 4.51(4.29-4.80) | <0.001 |
| Hemoglobin (g/L), (M, Q<sub>L</sub>-Q<sub>U</sub>) | 132 (126-138) | 128 (120-134) | <0.001 |
| Systolic blood pressure (mmHg), (M, Q<sub>L</sub>-Q<sub>U</sub>) | 120 (111-127) | 107 (100-115) | <0.001 |
| Diastolic blood pressure (mmHg), (M, Q<sub>L</sub>-Q<sub>U</sub>) | 80 (74-85) | 71 (65-77) | <0.001 |

Note: M: median, QL: lower quartile, Qu: upper quartile, HDP: hypertensive disorders of pregnancy.

### Associations between SCFAs and HDP

**Figure 1** shows the difference in plasma SCFAs levels between cases and controls. Amongst the seven SCFAs assayed, valerate was significantly lower in pregnant women with HDP than healthy controls (median level: 9 *vs*. 10 *ng*/ml, *P*=0.001). We failed to observe a statistically significant difference in the median level of other compounds such as acetate, propionate, isobutyrate, butyrate, isovalerate, and hexanoate (all *P*>0.05). A significant association between a higher level of valerate and a lower likelihood of HDP was also found by logistic regression (OR=0.679, 95% CI: 0.546-0.844, **Table 2**). This association persisted even after adjusting for maternal education level, BMI, systolic BP, hemoglobin, and fasting plasma glucose at the first antenatal visit (OR=0.699, 95% CI: 0.516-0.946). Notably, isobutyrate (OR=0.899, 95% CI: 0.826-0.980) and hexanoate (OR=0.686, 95% CI: 0.508-0.927) were also negatively associated with HDP, after adjustment for covariates.

**Figure 1.**
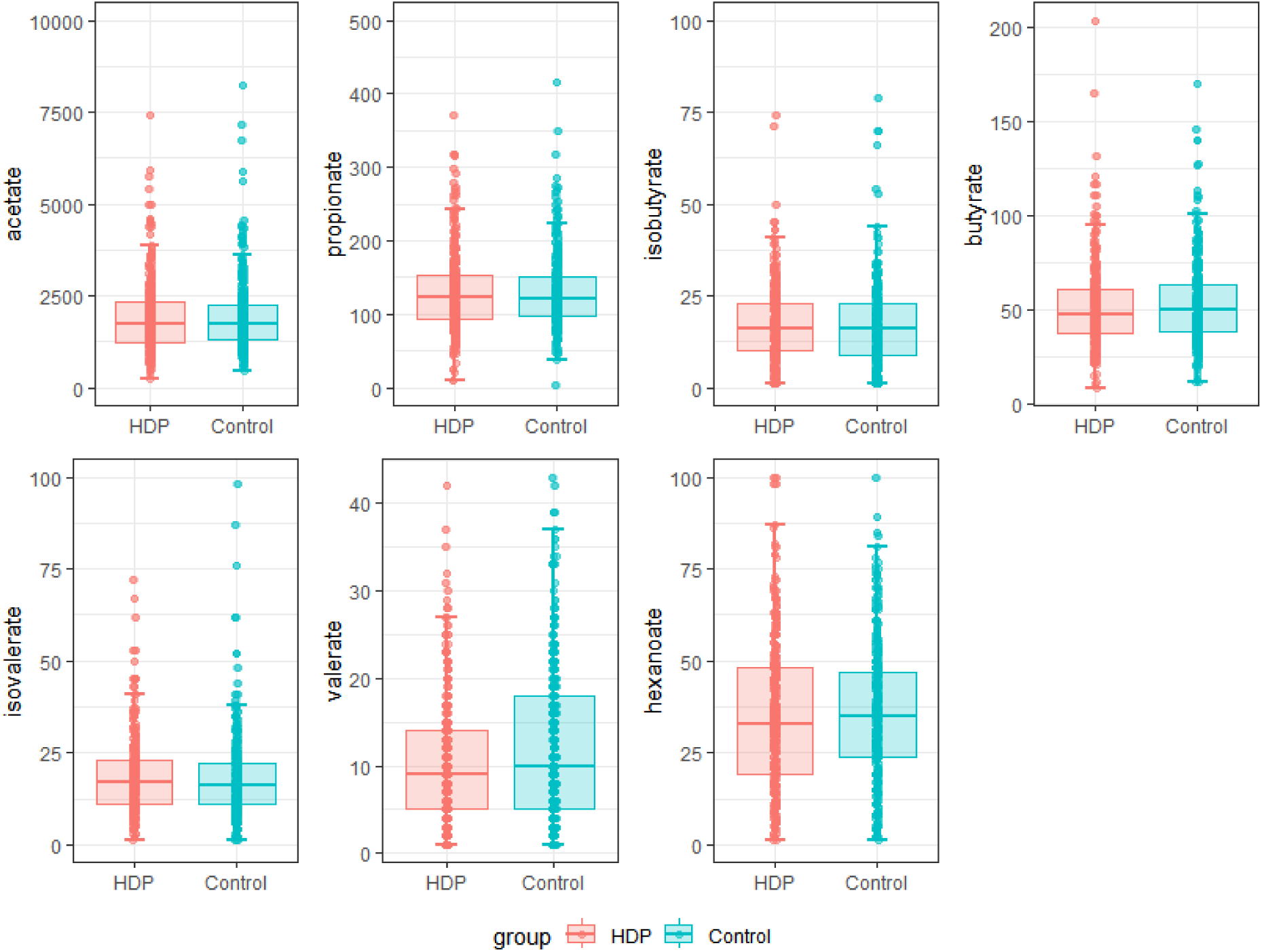
Distributions and comparisons of seven SCFAs between the case group and control group, Box plots showing the distributions and comparisons of SCFAs between the cases and controls. The box shows the median and interquartile range. Group differences in SCFAs levels were compared using the Wilcoxon signed-rank test.

**Table 2.** The associations of plasma SCFAs with HDP.

| SCFAs | Unadjusted |  | Adjusted* |  |
| --- | --- | --- | --- | --- |
|  | OR (95%CI) | P | OR (95%CI) | P |
| Acetate | 0.981(0.897-1.073) | 0.675 | 0.910(0.742-1.117) | 0.367 |
| Propionate | 1.048(0.919-1.195) | 0.484 | 0.938(0.772-1.139) | 0.517 |
| Butyrate | 0.911(0.778-1.065) | 0.242 | 0.831(0.667-1.034) | 0.097 |
| Isobutyrate | 0.975(0.915-1.039) | 0.438 | 0.899(0.826-0.980) | 0.015 |
| Isovalerate | 0.998(0.952-1.045) | 0.921 | 0.953(0.884-1.027) | 0.207 |
| Valerate | 0.679(0.546-0.844) | 0.001 | 0.699(0.516-0.946) | 0.020 |
| Hexanoate | 0.912(0.747-1.114) | 0.367 | 0.686(0.508-0.927) | 0.014 |
| Mixed exposure | 0.595(0.372-0.953) | 0.031 | 0.230(0.105-0.506) | 0.001 |

\*Adjusting for maternal education level, BMI, systolic BP, hemoglobin, and fasting plasma glucose at the first antenatal visit.
OR: odds ratio, CI: confidence interval; HDP: hypertensive of disorders in pregnancy; SCFAs: short chain fatty acids.

Although not all SCFAs showed a significant association with HDP, they were jointly associated with HDP (OR=0.595, 95% CI: 0.372-0.953), as suggested by the results of the WQS regression. This joint association is persistently significant after adjusting for conventional risk factors (OR=0.230, 95% CI: 0.105-0.506). The weight for each SCFA in the WQS regression is shown in **Supplementary Figure S2**. The top weight contribution was from hexanoate.

### Association between SCFAs and subtypes of HDP

In addition to HDP, plasma valerate was also significantly associated with subtypes of HDP such as GH (OR=0.706, 95% CI: 0.548-0.910) and PE/CH-PE (OR=0.609, 95% CI: 0.397-0.935), although these associations did not survive after adjustment for conventional risk factors (**Table 3**). Similarly, isobutyrate (OR=0.820, 95% CI: 0.690-0.973) and hexanoate (OR=0.508, 95% CI: 0.269-0.959) were also negatively associated with PE/CH-PE, after adjustment for covariates.

**Table 3.** The associations of plasma SCFAs with GH and PE/CH-PE.

| SCFAs | GH |  |  |  | PE/CH-PE |  |  |  |
| --- | --- | --- | --- | --- | --- | --- | --- | --- |
|  | Unadjusted |  | Adjusted* |  | Unadjusted |  | Adjusted* |  |
|  | OR(95%CI) | *P* | OR(95%CI) | *P* | OR(95%CI) | *P* | OR(95%CI) | *P* |
| Acetate | 0.988<br>(0.901-1.084) | 0.801 | 0.962<br>(0.772-1.199) | 0.729 | 0.92<br>1(0.697-1.216) | 0.561 | 0.763<br>(0.506-1.150) | 0.197 |
| Propionate | 1.099<br>(0.939-1.286) | 0.241 | 0.982<br>(0.761-1.268) | 0.889 | 0.831<br>(0.592-1.165) | 0.282 | 0.797<br>(0.512-1.242) | 0.316 |
| Butyrate | 0.965<br>(0.808-1.151) | 0.689 | 0.845<br>(0.646-1.106) | 0.220 | 0.737<br>(0.517-1.052) | 0.093 | 0.712<br>(0.456-1.112) | 0.136 |
| Isobutyrate | 0.982<br>(0.905-1.067) | 0.675 | 0.920<br>(0.805-1.051) | 0.219 | 0.965<br>(0.875-1.066) | 0.485 | 0.820<br>(0.690-0.973) | 0.023 |
| Isovalerate | 1.001<br>(0.949-1.056) | 0.961 | 0.967<br>(0.885-1.056) | 0.452 | 0.986<br>(0.896-1.085) | 0.774 | 0.878<br>(0.761-1.013) | 0.075 |
| Valerate | 0.706<br>(0.548-0.910) | 0.007 | 0.733<br>(0.495-1.086) | 0.121 | 0.609<br>(0.397-0.935) | 0.023 | 0.652<br>(0.395-1.075) | 0.094 |
| Hexanoate | 0.976<br>(0.777-1.225) | 0.833 | 0.724<br>(0.502-1.044) | 0.084 | 0.729<br>(0.477-1.114) | 0.144 | 0.508<br>(0.269-0.959) | 0.037 |
| Mixed exposure | 0.630<br>(0.422-0.890) | 0.032 | 0.740<br>(0.571-0.933) | 0.026 | 0.920<br>(0.680-1.740) | 0.527 | 0.725<br>(0.383-1.492) | 0.627 |

\*Adjusting for maternal education level, BMI, systolic BP, hemoglobin, and fasting plasma glucose at the first antenatal visit.
OR: odds ratio; CI: confidence interval; SCFAs: short chain fatty acids; GH: gestational hypertension; PE: preeclampsia; CH-PE: chronic hypertension with superimposed preeclampsia.

The seven SCFAs were also jointly associated with GH even after adjusting for conventional risk factors (OR=0.740, 95%CI: 0.571-0.933), but the top weight contribution was from valerate (**Supplementary Figure S3**). We failed to observe a significant joint association between SCFAs and PE/CH-PE.

### Results of MR analysis

In the literature, we successfully selected eligible IVs for acetate (13 SNPs), valerate (29 SNPs), isovalerate (35 SNPs), GH (20 SNPs), and PE (7 SNPs). The summarized data on these IVs are shown in **Supplementary Table S1**. Data on GH and PE included 8502 GH, 7212 PE patients and 194266 control pregnant women.

In the forward MR analysis, acetate, valerate, and isovalerate were considered as the exposure, GH and PE were considered as the outcome. The Cochran’s Q statistic and MR-Egger regression intercept suggested no heterogeneity and horizontal pleiotropy (all *P* > 0.05) (**Supplementary Table S2**). The IVW model with fixed effect showed that each standard deviation (SD) increment in genetically predicted plasma acetate (OR=0.784, 95% CI: 0.640-0.962, *P*=0.019), valerate (OR=0.575, 95% CI: 0.363-0.909, *P*=0.018) and isovalerate (OR=0.642, 95% CI: 0.428-0.963, *P*=0.032) were associated with a decreased risk of GH. Each SD increment in genetically predicted plasma acetate was also associated with a 25.4% reduction in the risk of PE (OR=0.746, 95% CI: 0.600-0.927, *P*=0.008). The leave-one-out sensitivity analysis showed that the causal effect estimated was not driven by any single SNP (**Figure 2**).

**Figure 2.**
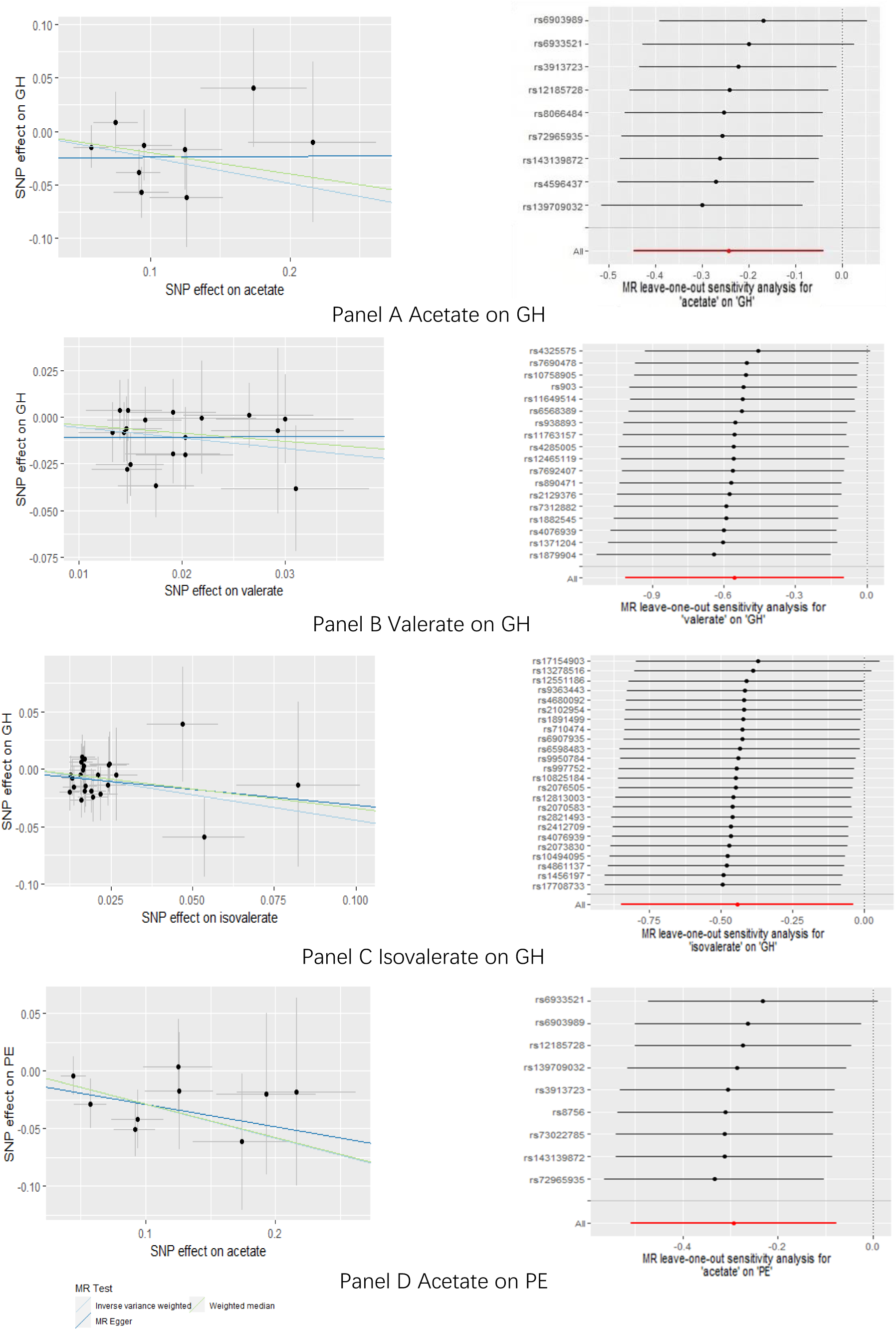
Scatter plots and forest plots of leave-one-out sensitivity analysis for genetic causal effect of SCFAs on GH/PE risk. The slopes of each line represent causal associations for each method in the scatter plot; In the forest plots of leave-one-out sensitivity analysis, each point represents the effect estimate for a genetic variant, expressed as a beta value, the lines extending from each point represent the 95% confidence intervals.

The reverse MR analysis did not reveal any significant associations between genetically determined risk of GH or PE and SCFAs (**Supplementary Table S3, Supplementary Figures S4-S5**).

## Discussion

Leveraging a nested case-control study in Chinese pregnant women, we for the first time found that plasma levels of three SCFAs such as valerate, isobutyrate, and hexanoate at 16-20 gw were significantly associated with a lower risk of future HDP during pregnancy, as well as its subtypes of GH and PE/CH-PE. Although not all SCFAs showed a significant individual association with HDP, they were jointly associated with HDP. These associations were also independent of conventional risk factors, e.g., obesity, glucose, and blood pressure at baseline. Further, a two-sample MR analysis demonstrated a significant association of genetically determined valerate and isovalerate with GH. These findings suggested that deficiency of SCFAs, valerate and isovalerate in particular, may precede and contribute to the development of HDP through mechanisms beyond these conventional factors.

In the nested case-control and MR analyses, the types of short-chain fatty acids associated with GH and PE were different, which could be due to differences in diet and genetic background between the two population. The SCFAs concentration depends on several factors, including diet, age, intestinal flora and genotype^17,18^. Meanwhile, diet and host genetics are important factors regulating the gut microbiota including SCFAs-producing bacteria^19–21^, and diet intervention can rapidly modify the composition of the microflora within a few days through direct and indirect mechanisms^22,23^. Compared with healthy pregnant women, patients with PE had lower levels of microbial diversity and SCFAs-producing bacteria ^24^. Chen et al ^25^reported PE patients had obvious dysbiosis with depleted beneficial bacteria including SCFAs-producing bacteria *Faecalibacterium* and *Akkermansia* and enriched opportunistic pathogens, moreover, the gut microbiome from patients with PE were transplanted to antibiotic-treated mice, which would provoke a dramatic, elevated pregestational blood pressure(BP) and a further increased BP after gestation in recipient mice. In additional, some studies found the low serum levels of acetate and butyrate were associated with preeclampsia development^26,27^. A study conducted by Jin et al^28^ demonstrated that the fecal, serum, and placental levels of propionic acid and butyric acid were significantly reduced in PE patients, and the abundance of *Akkermansia* which was negatively correlated with BP and urine protein was obviously decreased in patients with PE in contrast to normal late-pregnant women. Further oral administration of *Akkermansia muciniphila*, propionate, or butyrate to preeclamptic rats can significantly alleviated the symptoms of PE. Another study^12^ found the fecal levels of butyric and valeric acids were significantly decreased in patients with PE, and oral administration of butyrate to lipopolysaccharide(LPS) induced pregnant rats with hypertension could reduce the BP in these rats. However, not all the research findings are consistent. Li et al.^11^ reported the PE group had significantly higher levels of acetate, propionate, isobutyrate, and valerate than that in controls. The difference may be attributed to heterogeneity in subjects, regions and their diets. Currently, majority of researches focus on the relationship between SCFAs and PE, while studies on the relationship between SCFAs and GH is less reported.

SCFAs as the final significant metabolites of the gut microbiota plays an important role in host metabolism, immunity, and nutrition absorption^11^. The role of SCFAs in preventing pregnancy-induced hypertension may be related to its inhibitory effect on inflammation which plays an important role in the development of HDP^29–31^. SCFAs are sensed by specific G protein-coupled receptors (GPCRs) including GPR41/Ffar3 and GPR43/Ffar2, which are expressed in the gastrointestinal tract, adipose tissues, immune cells, and the autonomic nervous system, and regulate host energy homeostasis^32,33^. SCFAs specifically bind GPCRs, to inhibit the generation of T helper cells, thereby reducing the expression of nuclear factor-κB and the synthesis of pro-inflammatory factors such as interleukin-1β, interleukin-4, interleukin-5, interleukin-6 and tumor necrosis factor-α^34–36^. More importantly, by activating GPCRs, SCFAs can also increase the expression of regulatory T cells and the synthesis of anti-inflammatory factors γ interferon and interleukin-10 in the body^37,38^.

### Advantages and disadvantages

Existed research on SCFAs and HDP is insufficient, especially the study on the relationship between SCFAs and GH is limited. Our nested case-control study and MR analysis provide new evidence. In addition, MR is a powerful tool to assess causality, which can avoid biases commonly presented in observational studies. In our MR study, exposure data were extracted from the most recently published largest GWAS to detect causal effects. Furthermore, sensitivity analyses were performed to guarantee the satisfaction of model assumptions.

Of course, our study has some limitations. For example, the nested case-control study is hospital-based, and dietary information or fecal samples for each participant were not collected, and therefore could not explore the complete association among dietary intake, gut microbiome and circulating metabolites. Secondly, the research participants come from multiple hospitals in one region, and the extrapolation of the results to other regions is unknown. Thirdly, in MR study, only three types of SCFAs datasets were obtained in GWAS, and all GWAS data came from European population. Whether findings in our study would be consistent in other population remained to be investigated.

### Conclusions

In general, our nested case-control study confirms that plasma valerate level is a protective factor for HDP. And SCFAs co-exposure also can reduce risk of HDP. Besides, the large sample MR analysis indicates that plasma acetate, valerate and isovalerate may be causally negatively associated with a risk of GH, and acetate causally associated with a lower risk of PE. Therefore, our study suggests that it is necessary to appropriately increase the level of SCFAs during pregnancy to reduce the risk of HDP.

## Data Availability

The datasets used in the manuscript, code book and analytic code are available from the corresponding author on a reasonable request

## Acknowledgments

We gratefully acknowledge all participants of this study and thank the clinical staff at all participating hospitals for their support and contribution to this project.

## Authors’ contributions

Hao Peng, Lei Wu and Hongme Li designed research; Pei Feng, Yexiu Sun, Di Wu and He Xu conducted research; Dandan Wang and Pei Feng analyzed data; and Dandan Wang, Hao Peng and Hongmei Li wrote the paper. Hongmei Li had primary responsibility for final content. All authors read and approved the final manuscript.

## Ethics approval and consent to participate

The protocols of this study were approved by the Ethics Committee of Soochow University (Approval NO. ECSU-2019000118). All participants provided written informed consent to participate.

## References

1. Xiong T, Mu Y, Liang J, Zhu J, Li X, Li J, Liu Z, Qu Y, Wang Y, Mu D. Hypertensive disorders in pregnancy and stillbirth rates: a facility-based study in China. Bull World Health Organ. 2018;96:531–539. doi: 10.2471/BLT.18.208447

2. Roberts CL, Bell JC, Ford JB, Hadfield RM, Algert CS, Morris JM. The accuracy of reporting of the hypertensive disorders of pregnancy in population health data. Hypertens Pregnancy. 2008;27:285–297. doi: 10.1080/10641950701826695

3. Aronow WS. Hypertensive disorders in pregnancy. Ann Transl Med. 2017;5:266. doi: 10.21037/atm.2017.03.104

4. Umesawa M, Kobashi G. Epidemiology of hypertensive disorders in pregnancy: prevalence, risk factors, predictors and prognosis. Hypertens Res. 2017;40:213–220. doi: 10.1038/hr.2016.126

5. Brown MA, Magee LA, Kenny LC, Karumanchi SA, McCarthy FP, Saito S, Hall DR, Warren CE, Adoyi G, Ishaku S, et al. Hypertensive Disorders of Pregnancy: ISSHP Classification, Diagnosis, and Management Recommendations for International Practice. Hypertension. 2018;72:24–43. doi: 10.1161/HYPERTENSIONAHA.117.10803

6. Hutten JWM, Kuijpers JC, Velzen Dv, Wallenburg HCS. Pathogenesis of Pregnancy-Induced Hypertensive Disorders A Review of Experimental Animal Models. Clinical and Experimental Hypertension Part B: Hypertension in Pregnancy. 2009;10:287–306. doi: 10.3109/10641959109012916

7. Koren O, Goodrich JK, Cullender TC, Spor A, Laitinen K, Backhed HK, Gonzalez A, Werner JJ, Angenent LT, Knight R, et al. Host remodeling of the gut microbiome and metabolic changes during pregnancy. Cell. 2012;150:470–480. doi: 10.1016/j.cell.2012.07.008

8. Xu J, Moore BN, Pluznick JL. Short-Chain Fatty Acid Receptors and Blood Pressure Regulation: Council on Hypertension Mid-Career Award for Research Excellence 2021. Hypertension. 2022;79:2127–2137. doi: 10.1161/HYPERTENSIONAHA.122.18558

9. Yang F, Chen H, Gao Y, An N, Li X, Pan X, Yang X, Tian L, Sun J, Xiong X, et al. Gut microbiota-derived short-chain fatty acids and hypertension: Mechanism and treatment. Biomed Pharmacother. 2020;130:110503. doi: 10.1016/j.biopha.2020.110503

10. Yong W, Zhao Y, Jiang X, Li P. Sodium butyrate alleviates pre-eclampsia in pregnant rats by improving the gut microbiota and short-chain fatty acid metabolites production. J Appl Microbiol. 2022;132:1370–1383. doi: 10.1111/jam.15279

11. Li J, Wang L, Chen H, Yang Z, Chen S, Wang J, Zhou Y, Xuan R. The Diagnostic Potential of Gut Microbiota-Derived Short-Chain Fatty Acids in Preeclampsia. Front Pediatr. 2022;10:878924. doi: 10.3389/fped.2022.878924

12. Chang Y, Chen Y, Zhou Q, Wang C, Chen L, Di W, Zhang Y. Short-chain fatty acids accompanying changes in the gut microbiome contribute to the development of hypertension in patients with preeclampsia. Clin Sci (Lond). 2020;134:289–302. doi: 10.1042/CS20191253

13. Zhang S, Wang H, Zhu MJ. A sensitive GC/MS detection method for analyzing microbial metabolites short chain fatty acids in fecal and serum samples. Talanta. 2019;196:249–254. doi: 10.1016/j.talanta.2018.12.049

14. Kettunen J, Demirkan A, Wurtz P, Draisma HH, Haller T, Rawal R, Vaarhorst A, Kangas AJ, Lyytikainen LP, Pirinen M, et al. Genome-wide study for circulating metabolites identifies 62 loci and reveals novel systemic effects of LPA. Nat Commun. 2016;7:11122. doi: 10.1038/ncomms11122

15. Shin SY, Fauman EB, Petersen AK, Krumsiek J, Santos R, Huang J, Arnold M, Erte I, Forgetta V, Yang TP, et al. An atlas of genetic influences on human blood metabolites. Nat Genet. 2014;46:543–550. doi: 10.1038/ng.2982

16. Bowden J, Davey Smith G, Haycock PC, Burgess S. Consistent Estimation in Mendelian Randomization with Some Invalid Instruments Using a Weighted Median Estimator. Genet Epidemiol. 2016;40:304–314. doi: 10.1002/gepi.21965

17. Zietek M, Celewicz Z, Szczuko M. Short-Chain Fatty Acids, Maternal Microbiota and Metabolism in Pregnancy. Nutrients. 2021;13:1244. doi: 10.3390/nu13041244

18. Poll BG, Cheema MU, Pluznick JL. Gut Microbial Metabolites and Blood Pressure Regulation: Focus on SCFAs and TMAO. Physiology (Bethesda). 2020;35:275–284. doi: 10.1152/physiol.00004.2020

19. Amamoto R, Shimamoto K, Suwa T, Park S, Matsumoto H, Shimizu K, Katto M, Makino H, Matsubara S, Aoyagi Y. Relationships between dietary diversity and gut microbial diversity in the elderly. Benef Microbes. 2022;13:453–464. doi: 10.3920/bm2022.0054

20. Khachatryan ZA, Ktsoyan ZA, Manukyan GP, Kelly D, Ghazaryan KA, Aminov RI. Predominant role of host genetics in controlling the composition of gut microbiota. PLoS One. 2008;3:e3064. doi: 10.1371/journal.pone.0003064

21. Alvernaz SA, Wenzel ES, Nagelli U, Pezley LB, LaBomascus B, Gilbert JA, Maki PM, Tussing-Humphreys L, Peñalver Bernabé B. Inflammatory Dietary Potential Is Associated with Vitamin Depletion and Gut Microbial Dysbiosis in Early Pregnancy. Nutrients. 2024;16:935. doi: 10.3390/nu16070935

22. Wu N, Zhou J, Mo H, Mu Q, Su H, Li M, Yu Y, Liu A, Zhang Q, Xu J, et al. The Gut Microbial Signature of Gestational Diabetes Mellitus and the Association With Diet Intervention. Front Cell Infect Microbiol. 2021;11:800865. doi: 10.3389/fcimb.2021.800865

23. Ponzo V, Fedele D, Goitre I, Leone F, Lezo A, Monzeglio C, Finocchiaro C, Ghigo E, Bo S. Diet-Gut Microbiota Interactions and Gestational Diabetes Mellitus (GDM). Nutrients. 2019;11:330. doi: 10.3390/nu11020330

24. Liu J, Yang H, Yin Z, Jiang X, Zhong H, Qiu D, Zhu F, Li R. Remodeling of the gut microbiota and structural shifts in Preeclampsia patients in South China. Eur J Clin Microbiol Infect Dis. 2017;36:713–719. doi: 10.1007/s10096-016-2853-z

25. Chen X, Li P, Liu M, Zheng H, He Y, Chen MX, Tang W, Yue X, Huang Y, Zhuang L, et al. Gut dysbiosis induces the development of pre-eclampsia through bacterial translocation. Gut. 2020;69:513–522. doi: 10.1136/gutjnl-2019-319101

26. Hu M, Eviston D, Hsu P, Marino E, Chidgey A, Santner-Nanan B, Wong K, Richards JL, Yap YA, Collier F, et al. Decreased maternal serum acetate and impaired fetal thymic and regulatory T cell development in preeclampsia. Nat Commun. 2019;10:3031. doi: 10.1038/s41467-019-10703-1

27. Altemani F, Barrett HL, Gomez-Arango L, Josh P, David McIntyre H, Callaway LK, Morrison M, Tyson GW, Dekker Nitert M. Pregnant women who develop preeclampsia have lower abundance of the butyrate-producer Coprococcus in their gut microbiota. Pregnancy Hypertens. 2021;23:211–219. doi: 10.1016/j.preghy.2021.01.002

28. Jin J, Gao L, Zou X, Zhang Y, Zheng Z, Zhang X, Li J, Tian Z, Wang X, Gu J, et al. Gut Dysbiosis Promotes Preeclampsia by Regulating Macrophages and Trophoblasts. Circ Res. 2022;131:492–506. doi: 10.1161/CIRCRESAHA.122.320771

29. Salazar Garcia MD, Mobley Y, Henson J, Davies M, Skariah A, Dambaeva S, Gilman-Sachs A, Beaman K, Lampley C, Kwak-Kim J. Early pregnancy immune biomarkers in peripheral blood may predict preeclampsia. J Reprod Immunol. 2018;125:25–31. doi: 10.1016/j.jri.2017.10.048

30. Socha MW, Malinowski B, Puk O, Dubiel M, Wicinski M. The NLRP3 Inflammasome Role in the Pathogenesis of Pregnancy Induced Hypertension and Preeclampsia. Cells. 2020;9:1–13. doi: 10.3390/cells9071642

31. Gomez-Lopez N, Motomura K, Miller D, Garcia-Flores V, Galaz J, Romero R. Inflammasomes: Their Role in Normal and Complicated Pregnancies. J Immunol. 2019;203:2757–2769. doi: 10.4049/jimmunol.1900901

32. Miyamoto J, Kasubuchi M, Nakajima A, Irie J, Itoh H, Kimura I. The role of short-chain fatty acid on blood pressure regulation. Curr Opin Nephrol Hypertens. 2016;25:379–383. doi: 10.1097/MNH.0000000000000246

33. Brown AJ, Goldsworthy SM, Barnes AA, Eilert MM, Tcheang L, Daniels D, Muir AI, Wigglesworth MJ, Kinghorn I, Fraser NJ, et al. The Orphan G protein-coupled receptors GPR41 and GPR43 are activated by propionate and other short chain carboxylic acids. J Biol Chem. 2003;278:11312–11319. doi: 10.1074/jbc.M211609200

34. Sam QH, Ling H, Yew WS, Tan Z, Ravikumar S, Chang MW, Chai LYA. The Divergent Immunomodulatory Effects of Short Chain Fatty Acids and Medium Chain Fatty Acids. Int J Mol Sci. 2021;22:6453–6466. doi: 10.3390/ijms22126453

35. Li M, van Esch B, Wagenaar GTM, Garssen J, Folkerts G, Henricks PAJ. Pro- and anti-inflammatory effects of short chain fatty acids on immune and endothelial cells. Eur J Pharmacol. 2018;831:52–59. doi: 10.1016/j.ejphar.2018.05.003

36. Nakajima A, Nakatani A, Hasegawa S, Irie J, Ozawa K, Tsujimoto G, Suganami T, Itoh H, Kimura I. The short chain fatty acid receptor GPR43 regulates inflammatory signals in adipose tissue M2-type macrophages. PLoS One. 2017;12:1–18. doi: 10.1371/journal.pone.0179696

37. Kaye DM, Shihata WA, Jama HA, Tsyganov K, Ziemann M, Kiriazis H, Horlock D, Vijay A, Giam B, Vinh A, et al. Deficiency of Prebiotic Fiber and Insufficient Signaling Through Gut Metabolite-Sensing Receptors Leads to Cardiovascular Disease. Circulation. 2020;141:1393–1403. doi: 10.1161/CIRCULATIONAHA.119.043081

38. Correa RO, Vieira A, Sernaglia EM, Lancellotti M, Vieira AT, Avila-Campos MJ, Rodrigues HG, Vinolo MAR. Bacterial short-chain fatty acid metabolites modulate the inflammatory response against infectious bacteria. Cell Microbiol. 2017;19:e12720. doi: 10.1111/cmi.12720

